# Delay and bias in PubMed medical subject heading (MeSH^®^) indexing of respiratory journals

**DOI:** 10.1101/2020.10.01.20205476

**Authors:** Ruth M. Hadfield

## Abstract

**Introduction:** PubMed is a primary global open-access literature research database. Articles on PubMed are indexed manually with medical subject headings (MeSH^®^) to facilitate more complete literature searches. We aimed to determine the length of delay from publication to MeSH^®^ indexing for key respiratory journals and to investigate whether delays are increasing over time and whether there are country or impact-factor specific biases in indexing.

**Methods:** PubMed was searched for the keyword ‘asthma’ for the 10 year period June 2009 to June 2019. MEDLINE fields including journal title, publication date, PubMed entrez date and MeSH^®^ indexing date were extracted and delay in indexing was calculated in days.

**Results:** Twenty-nine respiratory journals had delay from publication to MeSH^®^ indexing ranging from an average of 153.6 days to 409.9 days; 5/29 (17.2%) had never been indexed for PubMed. There was a significantly longer delay in MeSH^®^ indexing for UK-based publications (mean delay of 281.7 days) compared to USA-based publications (mean delay of 214.9 days; mean difference 66.8 days, 95% CI 60.8, 72.8, P < 0.0001). Delays in MeSH^®^ indexing increased over time.

**Conclusion:** There are long and increasing delays in PubMed MeSH^®^ indexing of respiratory journals. All PubMed users should be aware that systematic literature searches that rely on MeSH^®^ search terms or utilise PubMed filters are likely to exclude recent research and citations from key journals. Researchers and clinicians need to be aware of these delays and biases to ensure their literature searches are both up-to-date and complete.

## Introduction

PubMed was established by the US National Library of Medicine (NLM) and is a primary global open-access literature research database. For many clinicians and researchers who do not have access to subscription-based databases such as Ovid^®^ (Wolters Kluwer) and Embase^®^ (Elsevier), literature research is often primarily conducted in PubMed.

Medical Subject Heading (MeSH^®^) indexing is a time-consuming manual process in which degree-qualified indexers who are specialists in the subject matter read the full manuscript, identify important topics, determine subject content, and assign appropriate headings and sub-headings.[1] MeSH^®^ headings facilitate the expansion and refinement of literature searches and ensure completeness; for example when a study is a randomised controlled trial but these words have not been used in the title or abstract, a MeSH^®^ indexer will add these terms and also add the term ‘clinical trial’ to the ‘study type’ field.

Most high ranked journals are entered into the PubMed database when they are accepted for publication however as MeSH^®^ indexing is a manual task there is a lag time. This delay has been investigated by two studies of pharmacy journals with one reporting an average of 177 days[2] for articles published in 2012 and the other a median of 52 to 252 days for articles published in 2012-2013.[3] A third study reported that the number of articles not MeSH^®^ indexed increased from 8% in 2008 to 34% in 2017.[4]

Developers of machine learning algorithms that will utilise the PubMed database have also identified that time-sensitive features, such as MeSH^®^ indexing and study type, have a detrimental effect on the completeness of a literature search.[5]

We conducted this study to investigate the delay in MeSH^®^ indexing of asthma articles in key respiratory journals. Our primary aim was to determine the length of delay from publication to indexing and whether delays are increasing over time. Secondary aims were to investigate whether there was preferential indexing for high-ranking journals, or journals published in specific countries.

## Methods

PubMed (NCBI, PubMed.gov, US National Library of Medicine, National Institutes of Health, Bethesda, MD, USA) was searched for the keyword ‘asthma’ for the 10 year period June 2009 to June 2019. The search string used is available in Appendix A. The full MEDLINE record was downloaded for each publication and data was filtered to include the journal title (JT), publication date, PubMed entrez date (EDAT; date entered into PubMed database) and MeSH^®^ indexing date (MHDA). The difference between the entrez date and the MeSH^®^ indexing date was calculated in days.

Journals were tabulated according to rank in the SciMago Pulmonary and Respiratory journal listing (Table 1). https://www.scimagojr.com/journalrank.php?category=2740 [Accessed 17 June 2019]

**Table 1.**
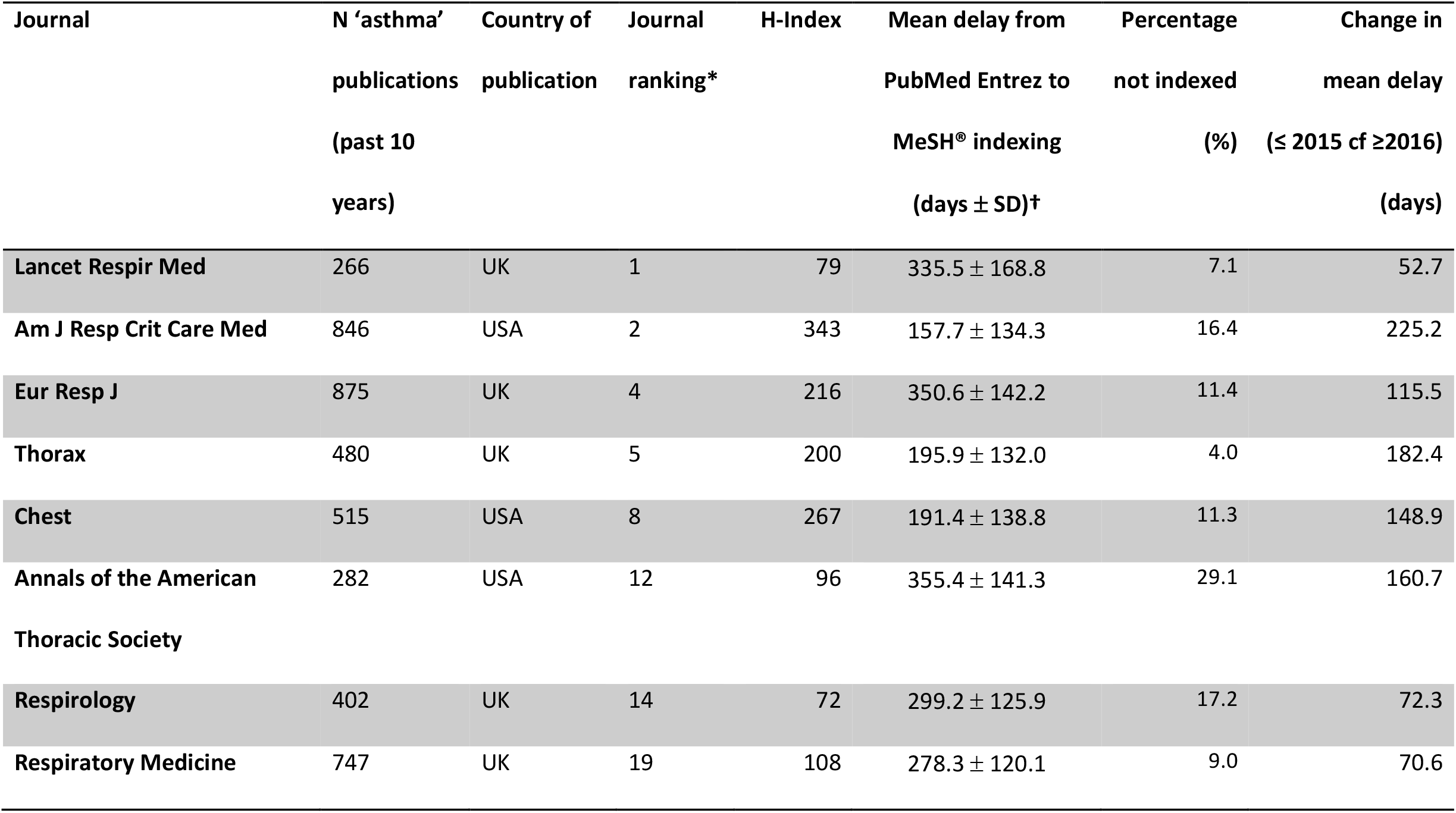
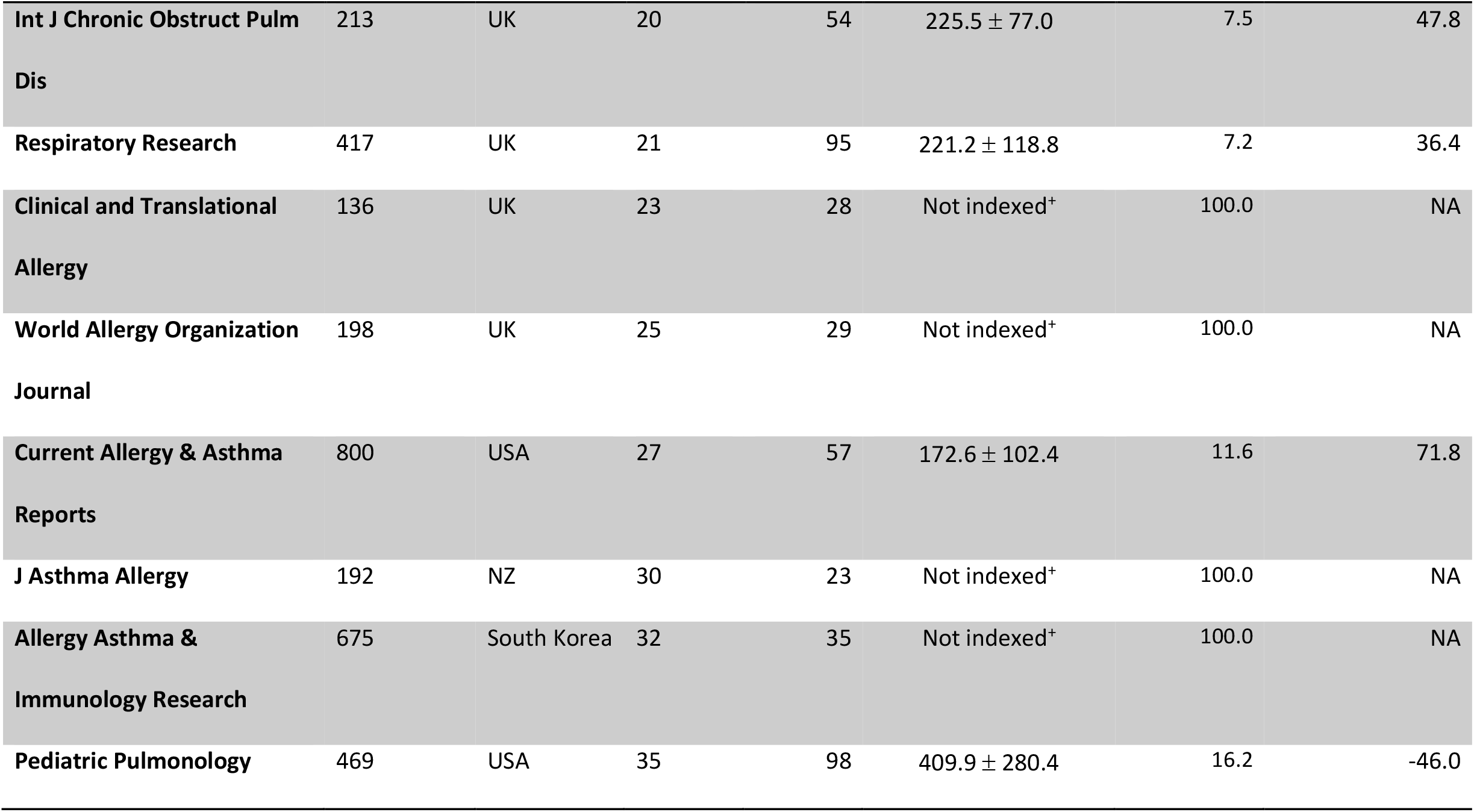
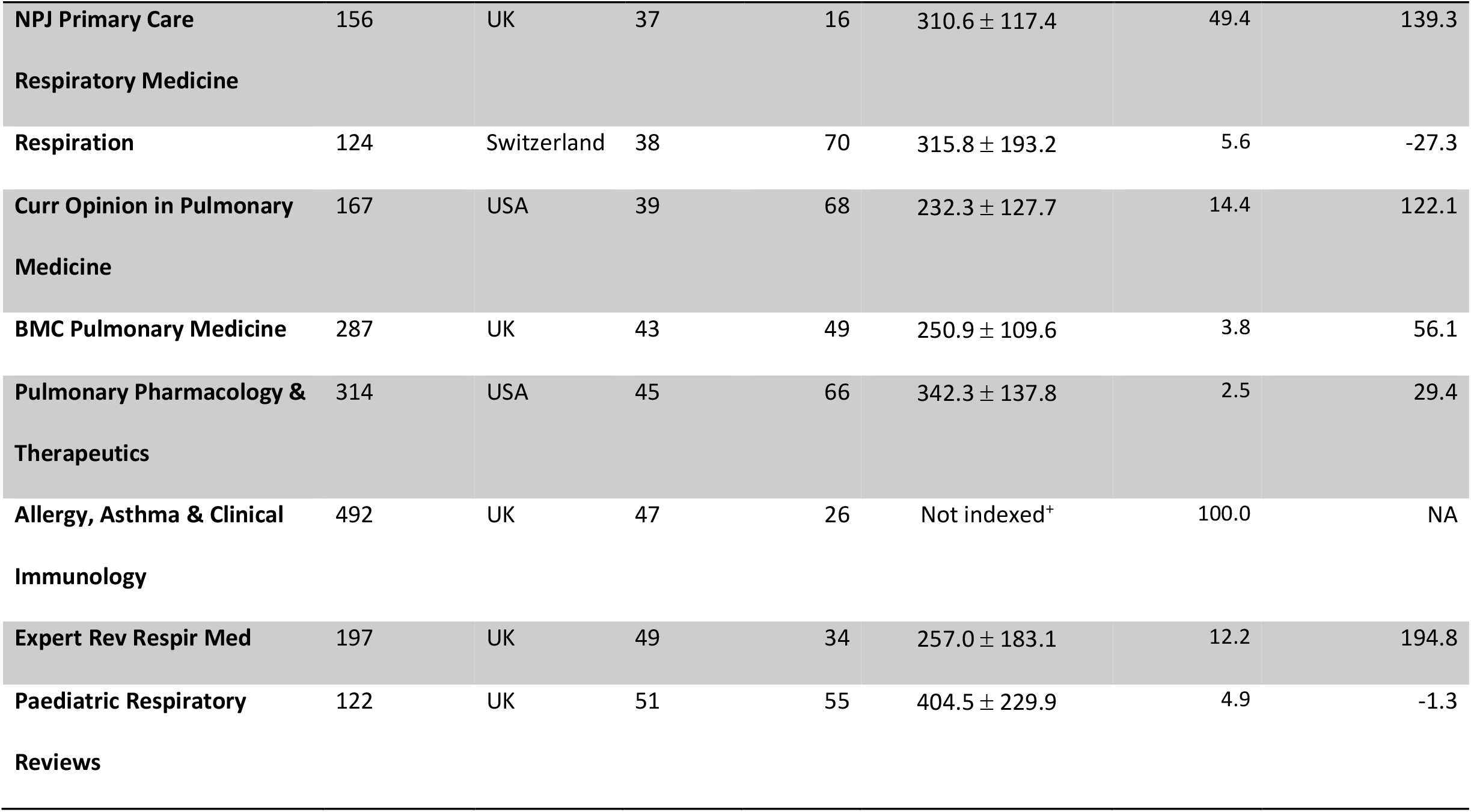
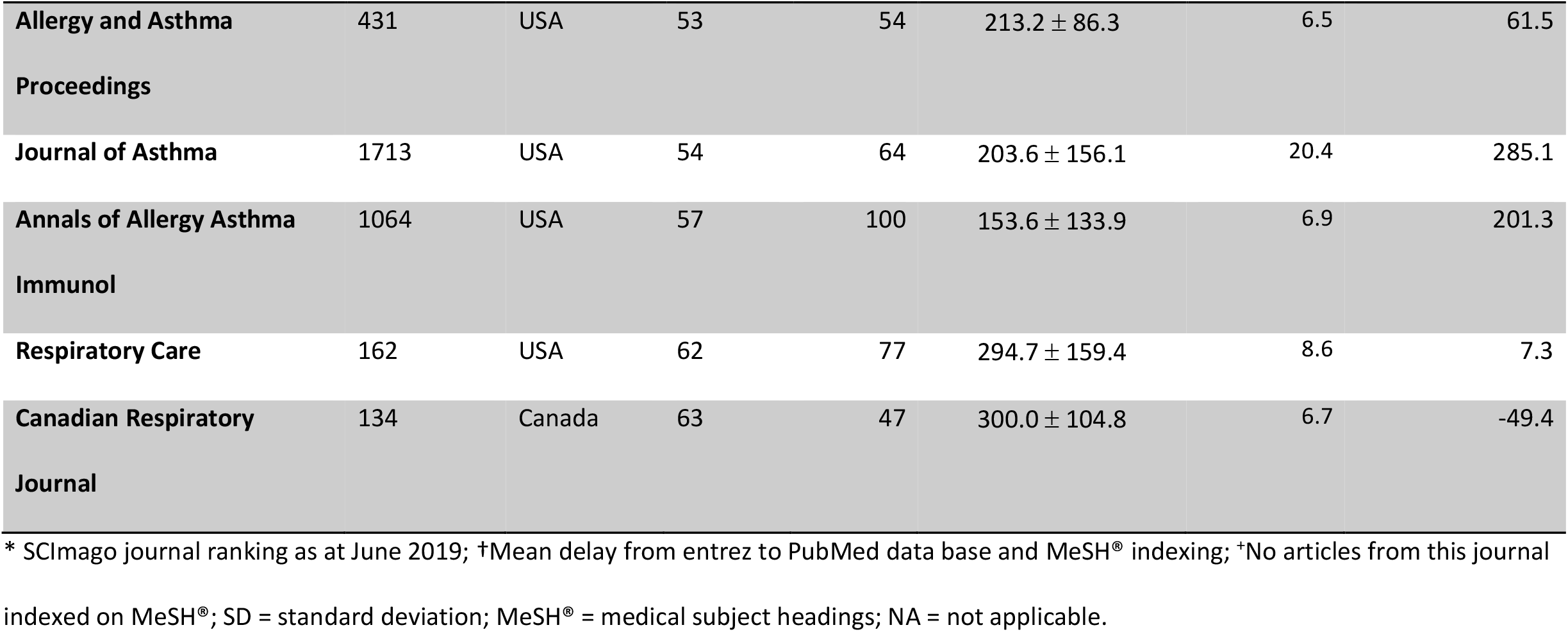
Respiratory journals and key metrics.

Journal titles focusing on primarily clinical research were selected from the top 100 respiratory journals. Journals that had <100 publications on asthma in the 10 year period were excluded as were those focused on lung cancer, surgery or laboratory research. Excluded journals due to low numbers of asthma articles were: The Journal of heart and lung transplantation: the official publication of the International Society for Heart Transplantation (n=1); Sleep Medicine Reviews (n=4); Journal of Thoracic Oncology (n=1); European Respiratory Review : an official journal of the European Respiratory Society (n=60); BMJ Open Respiratory Research (n=49); Asthma Research and Practice (n=49); Clinics in Chest Medicine (n=57); ERJ Open Research (n=63);COPD journal of Chronic Obstructive Pulmonary Disease (n=70); Therapeutic Advances in Respiratory Disease (n=73); Journal of Aerosol Medicine and Pulmonary Drug Delivery (n=85); Chronic Respiratory Disease (n=46); Seminars in respiratory and critical care medicine (n=43); Breathe (Sheffield, England) (n=34); Journal of Breath Research (n=66).

Results were analysed using Microsoft Excel software. Means were compared using the students T-test.

## Results

Data were collected for 29 different respiratory journals. The average delay from the date of first entry (‘Entrez’) into the PubMed database to the date of MeSH^®^ indexing ranged from 153.6 days to 409.9 days for the selected journals (Table 1; Figure 1).

**Figure 1.**
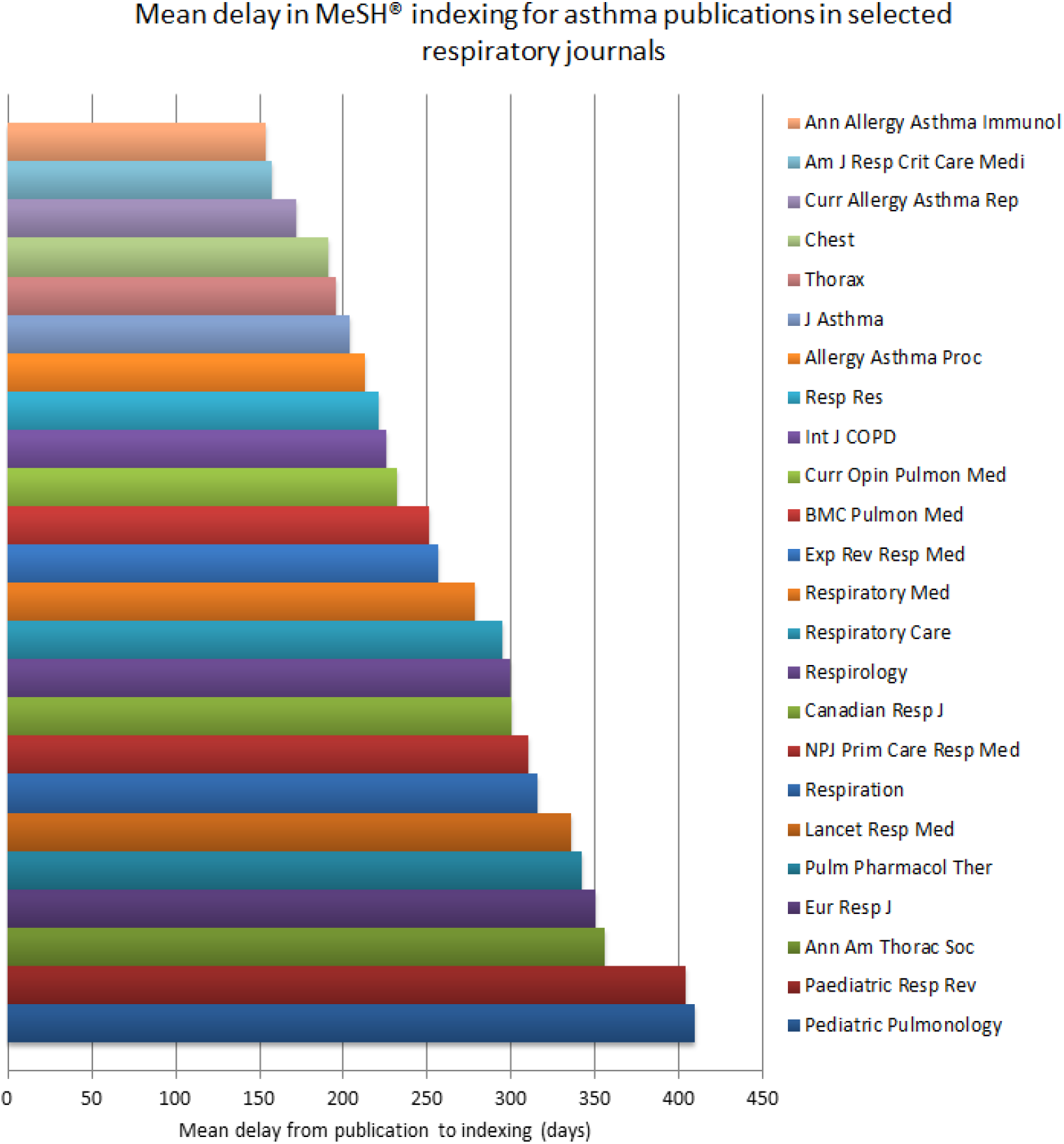
Graph showing the mean delay from publication (Entrez to PubMed) to MeSH^®^ indexing for asthma articles in selected respiratory journals.

Of the journals analysed 5 (17.2%) had never been indexed for PubMed. Of the remaining 24 journals the percentage of non-indexed articles ranged from 2.5% to 49.4% (Table 1). All US-based journals were indexed (11/11; 100% indexed) compared to only a proportion of non-US-based journals (13/18; 72.2% indexed; Table 1). Journals not indexed were based in the UK (n=3), South Korea (n=1) and New Zealand (n=1).

There was a significantly longer delay in MeSH^®^ indexing for UK-based publications with a mean delay of 281.7 days compared to US-based publications with a mean delay of 214.9 days (mean difference 66.8 days; 95% CI 60.8, 72.8; P < 0.0001).

For journals indexed, 20/24 (83%) had experienced increased delay in indexing, with higher mean delays for articles published since 2016 compared to those published between 2010-2015 (Table 1; Figure 1). There was no correlation between H-Index and delay in indexing (r^2^ = 0.0978).

## Discussion

PubMed is a primary resource for searching the medical literature and widely used by researchers and clinicians. It is also a key tool for those outside academic institutions who do not have access to subscription-based bibliographic databases for example primary healthcare and allied health professionals, and non-government organisations.[6]

Our findings highlight the lengthy and increasing delays in PubMed MeSH^®^ indexing. Furthermore a considerable proportion (17.2%) of key, high-impact respiratory journals are not MeSH^®^ indexed at all.

Reasons for not being indexed are likely to be multifactorial. The US NLM lists criteria for journal inclusion on its website[7] and the Literature Selection Technical Review Committee (LSTRC) meets three times per year to review and assess quality of journal title. Journal titles must meet a number of criteria in order to be indexed including providing XML-tagged data, access to content and permanent preservation facilities.

Although skilled literature searchers are likely to be aware that MeSH^®^ terms alone cannot be used reliably for an up-to-date literature search, many clinicians may be unaware of this. Of greater concern is that many of the available filters on the left hand panel of the PubMed search screen (Figure 2) rely on indexing terms. For example if a PubMed literature search for the keyword ‘asthma’ is carried out and then the ‘Article type’ filter ‘Clinical trial’ is selected, only MeSH^®^ indexed articles will appear in the search results and all non-indexed articles will be excluded. The ‘Article type’ field is added at the time of indexing, other fields that are dependent on indexing include ‘Language’, ‘Species’, and ‘Ages’.

**Figure 2.**
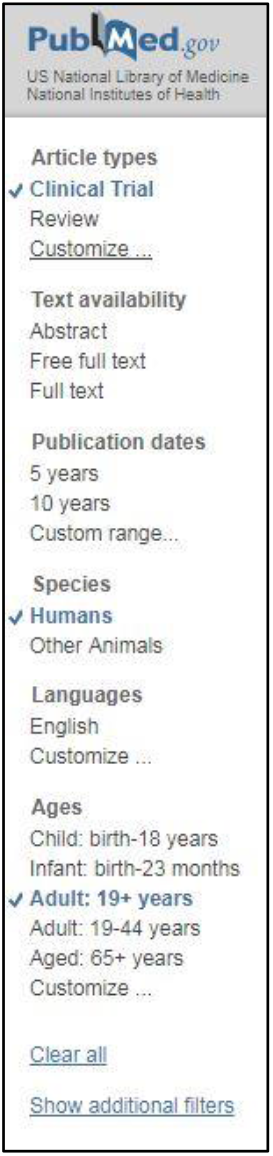
The PubMed ‘filter’ panel allows selection of a subset within a literature search. For example ‘Article types’, ‘Species’ and ‘Ages’ all rely on fields added at the time of MeSH^®^ indexing. If an article has not been MeSH^®^ indexed it will be excluded from the search if these filters are applied.

Conversely, omitting MeSH^®^ is not advisable as indexers add in additional asthma papers that do not contain the term ‘asthma’ in the title, abstract or keywords. For instance, in the past 5 years, 1,537 articles were added by indexers to the MeSH^®^ heading ‘asthma’ even though the term ‘asthma’ was not in the title or abstract. It is therefore advisable to use both the keyword and the MeSH^®^ heading with a boolean ‘OR’ in the search string; filters should not be used for an up-to-date search.

As the number of medical publications has grown over so time so too has the task of indexing.[8] Indeed the US NLM report that the number of citations indexed for MEDLINE^®^ has nearly doubled in the past 15 years (Figure 3). In our study for most respiratory journals (83%) there was an increase in delay in indexing over time. There may have also been a delaying effect of the most recent US government shutdown periods which occurred in January 2018 and December 2018/January 2019 and affected the operations of the NIH and NLM.

**Figure 3.**
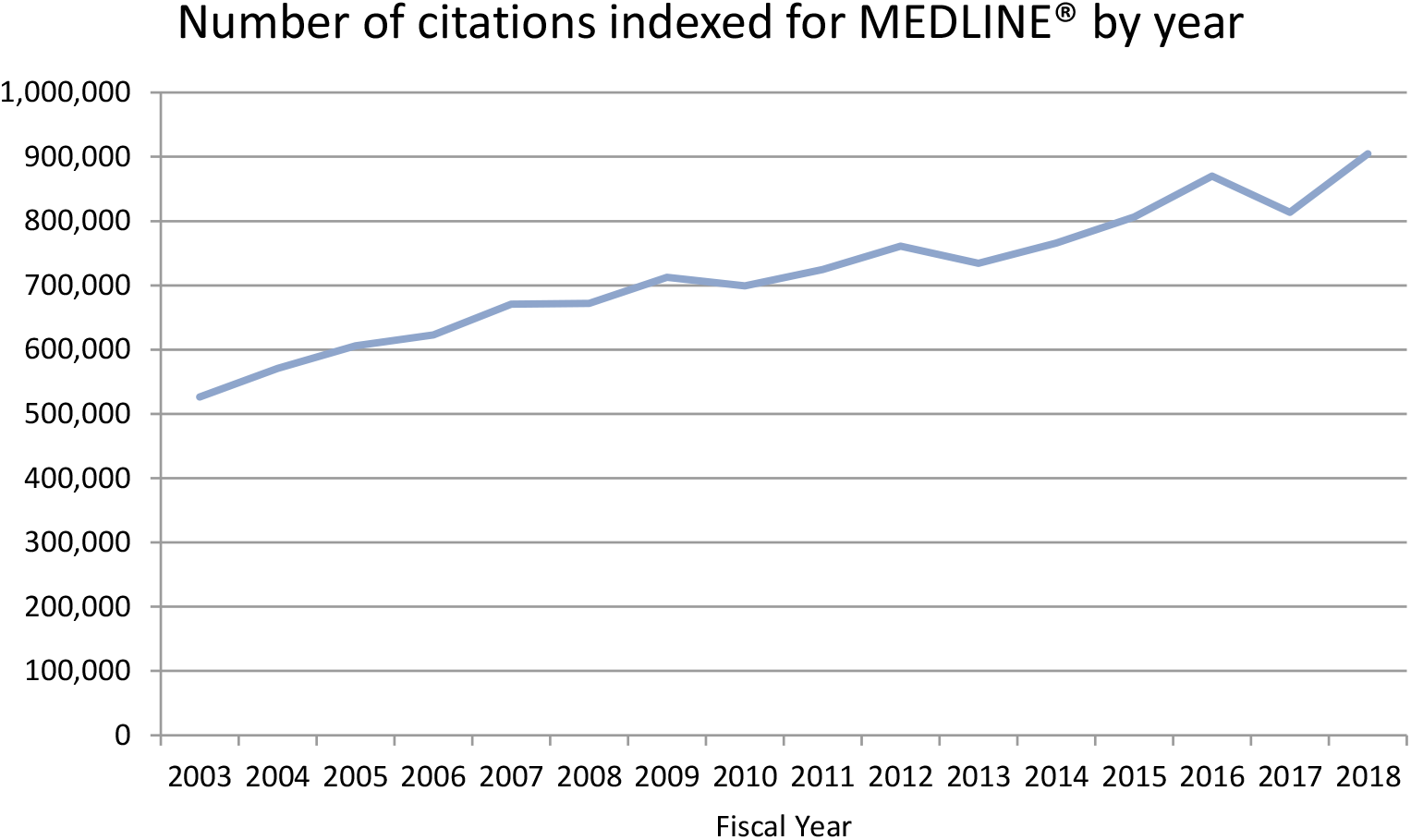
The number of citations indexed for MEDLINE^®^ each year [Data from NLM website URL https://www.nlm.nih.gov/bsd/bsd_key.html]

While it appears there is no preferential indexing of journals with high impact factors, respiratory journals that are US-based were indexed significantly faster than those that are UK-based.

We can conclude that systematic literature searches that rely on MeSH^®^ search terms alone or PubMed filters are likely to be out-of-date and possibly inherently biased. Researchers and clinicians need to be aware of these delays and biases to ensure their literature searches are as up to date and complete as possible. This is particularly relevant for the development of search strings for systematic literature reviews. It is also important that artificial intelligence and machine learning developers are aware that searches relying solely on MeSH^®^ index may be out of date and introduce bias.[9]

## Data Availability

All data is available from the author on request

## Summary table

### What was already known on the topic

- Substantial delay in PubMed/MEDLINE MeSH^®^ indexing has been reported by two studies with a third study observing increasing numbers of non-indexed publications.
- Machine-learning developers that utilise the PubMed database have identified that the time delay of MeSH^®^ indexing has a detrimental effect on the completeness of a literature search.

### What this study added to our knowledge

- Using the example of respiratory journals, this study found that there is a long delay in PubMed/MEDLINE MeSH^®^ indexing (range 153.6 days to 409.9 days).
- The delay in indexing has increased significantly over time.
- A large number of journals are never indexed (17.2%).
- There is bias in indexing with US-based respiratory journals indexed sooner than UK-based respiratory journals
- UK-based journals less likely to be indexed.

### Highlights

- There is a long delay in MeSH^®^ indexing and the delay is increasing over time
- Non-US-based respiratory journals are less likely to be indexed than US-based journals
- US-based respiratory journals are indexed sooner than UK-based respiratory journals
- No literature search should rely solely on MeSH^®^ index terms
- PubMed filters should be used with caution

## Declaration of interests

None.

## Appendix A

Search string: (“asthma”[MeSH Terms] OR “asthma”[All Fields]) AND “Insert full journal name here”[Journal] AND (“2009/06/19”[PDat] : “2019/06/16”[PDat])

## Notes

### Competing Interest Statement

The authors have declared no competing interest.

### Funding Statement

No external funding was received.

